# Incidence and Predictors of Antiretroviral Treatment Failure among Children in Public Health Facilities of Kolfe Keranyo Sub-City, Addis Ababa, Ethiopia: Institution-based retrospective cohort study

**DOI:** 10.1101/2022.03.24.22272879

**Authors:** Meseret Misasew, Daniel Angassa

## Abstract

**Background:** Human immunodeficiency virus (HIV) infection is a public health concern globally. The numbers of people living with HIV worldwide in 2018 was estimated at 37.9 million; from those, 1.7 million are children. Globally, 62% of the 37.9 million people were receiving Antiretroviral treatment (ART); 53% had achieved viral suppression. This study aimed to assess the incidence and predictors of Antiretroviral treatment failure among children in Kolfe Keranyo sub-city, Addis Ababa, Ethiopia.

**Methods:** An institution-based retrospective cohort study was conducted among 250 children who were enrolled to first-line Antiretroviral treatment from January 2013 to May 2020 in Kolfe Keranyo sub-city. Data was collected by using data extraction checklist and data were extracted by reviewing children’s medical chart and electronic database. Kaplan–Meier method was used to estimate the probability of treatment failure. During bivariable analysis variables with p-value < 0.25 were taken for multivariable Cox regression analysis to assess predictors of treatment failure. Statistically significant association was declared at p-value < 0.05 with 95% confidence interval.

**Result:** The overall treatment failure rate within the follow-up period was 17.2%. This study also found that the overall incidence density rate was 3.45% (95% CI: 2.57-4.67) per 1000 person-month observation. Infant prophylaxis for PMTCT (AHR: 3.59, 95% CI: 1.65-7,82), drug substitution (AHR: 0.18, 95% CI: 0.09-0.37), AZT/3TC/NVP based regimen (AHR: 2.27, 95% CI: 1.14-4.25), and more than 3 episodes of poor ART adherence (AHR: 2.27, 95% CI: 1.17-4.38) were found to be predictors of treatment failure among children.

**Conclusion:** High proportion of treatment failure was found among children on first-line ART in Kolfe Keranyo sub-city, Addis Ababa. Infant prophylaxis for PMTCT, drug substitution, initial regimen, and poor ART adherence were found to be predictors of first-line ART treatment failure. Close follow-up of children on medication adherence and effective trainings to health care professionals need to be considered.

## Introduction

Human immunodeficiency virus (HIV) infection is a public health concern globally, especially in Sub-Saharan African countries. The numbers of people living with HIV worldwide in 2018 have estimated at 37.9 million out of these 1.7 million are children. Sub-Saharan Africa remains the region most heavily affected by HIV, accounted for 67.5% of HIV infections worldwide in adults and 90% in children (1–4). New HIV infection in 2018 account 1.7 million adults and 160,000 children and 54% of children living with HIV were receiving lifelong ART in low and middle-income countries (5).

Ethiopia, to reduce the problem of HIV, began providing Antiretroviral Treatment (ART) in 2003 and free ART was launched in 2005 (1,6). According to the 2017 Ministry of Health estimate, 722,248 Ethiopians are currently living with HIV but only 420,000 are taking ART. Although ART coverage for adults has reached 61%, it remains low for children (33%) (7). In Ethiopia there is a significant pediatric HIV-1 burden with approximately 63,227 infected children and estimated 5479 new HIV infected children with 3200 AIDS-related children deaths occurring annually (1,2).

Monitoring of ART has used to ensure successful treatment, identify adherence problems, and determine whether ART regimens should be switched in case of treatment failure (2,3). Treatment failure is a suboptimal response or a lack of sustained response to treatment (8,9). Treatment failure is measured by clinical, immunological and virological failures.

The global target of a sustainable developmental goal (SDG) is to achieve 90-90-90 treatment targets at the end of 2020. These targets are first 90% of people living with HIV to know their status, second 90% of those who know their status are to be on treatment, and third 90% of those on treatment are to be virally suppressed (2).

A systematic review and meta-analysis conducted in 2017 in Ethiopia depicted that the pooled prevalence of ART failure was estimated to 15.3% by using WHO treatment failures criteria (10). Children and adolescents are more likely to fail on ART than adults. A retrospective cohort study conducted at Addis Ababa in four hospitals, showed that 14.1% of treatment failure were in children (11).

Even though, various global and local studies indicated a high proportion of treatment failure among children (11–13), little is known about the predictors of virological failure among children in Ethiopia. Most studies define treatment failure based on the two WHO criteria, clinical and immunological criteria. Currently, routine viral load workup for monitoring treatment failure is being implemented in Ethiopia. Therefore, based on routine viral load measurement along with clinical and immunological criteria, this study aimed to assess the incidence and predictors of treatment failure among children on first line antiretroviral therapy in Kolfe Keranyo sub-city Addis Ababa, Ethiopia.

## Materials and Methods

### Study design, setting and population

An institution-based retrospective cohort study was conducted from January to March 2021 among children enrolled to first-line ART from January 2013 to May 2020 in selected health facilities in Kolfe Keranyo sub-city. Kolfe Keranyo sub city is one of the eleven sub-cities of Addis Ababa, the capital city of Ethiopia. Administratively, the sub-city is divided in to 15 Woredas encompassing 1 hospital and 11 functional governmental health centers. The total population of Kolfe Keranyo sub-city in 2019 was estimated to be around 576,443. The sub-city is providing ART service to 14,161 people living with HIV. All children aged less than 15 years old who started receiving ART and are on follow-up at least for six months at the selected health facilities in Kolfe Keranyo sub-city from 2013 and 2020 were included in this study. Whereas, children with incomplete medical records were excluded (14).

### Sample size and sampling technique

The required sample size was determined by using a single population proportion formula based on a similar study conducted among pediatric population in Ethiopia which showed a 17.3% prevalence of ART failure (15). An assumption of 5% margin of error, 95% confidence interval, and 80% of power were considered. Additionally, after considering a 10% non-response rate for incomplete medical records, the final sample size was 250.

The health facilities were selected based on their number of pediatric ART clients. Facilities with more than ten pediatric ART clients were selected. From the 11 health centers one hospital of in the sub-city, five health centers and one hospital were selected. After preparing a sampling frame, the total sample size was then proportionally allocated to the selected health facilities. Finally, medical charts were selected with a simple random sampling technique (**Fig. 1)**.

**Fig. 1:**
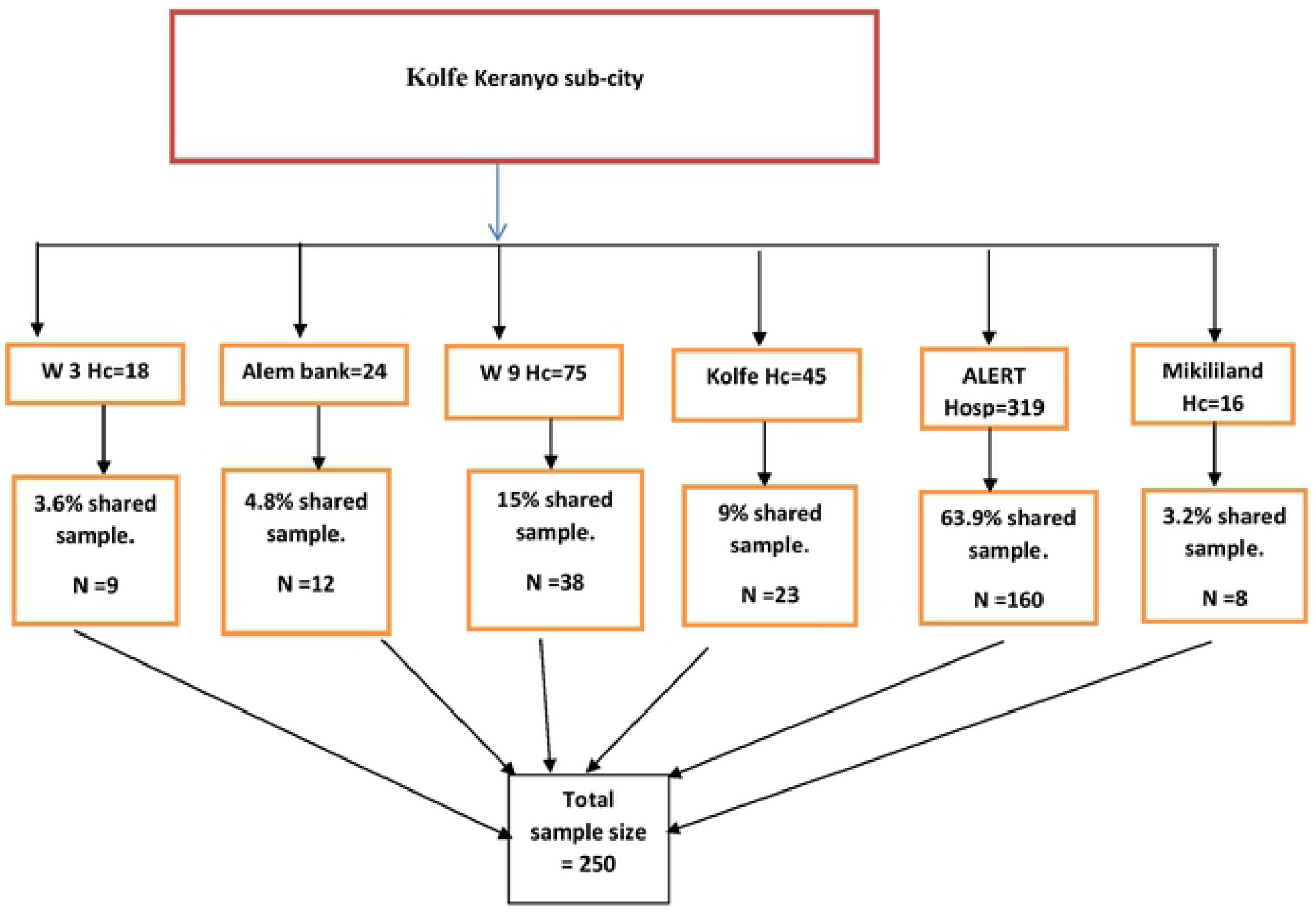
Sample size distribution of study participants in Kolfe Keranyo sub-city, Addis Abada, Ethiopia.

### Data collection and quality control

A structured data abstraction checklist in English language containing the most relevant variables regarding ART treatment outcomes was adopted from different previous studies. Data were collected both from patient follow up charts and electronic database. The data abstraction checklist included socio-demographic, ART, clinical, laboratory, and treatment related characteristics. The data was retrieved by eight data clerks who have diploma in information technology and experience in managing ART data in different facilities. One data collector was assigned to collect the data from each health center and three data collectors for the hospital. A one day of training on data collection tools and process was conducted. The adopted data collection checklist was pretested to check for its completeness and consistency. The principal investigator and the assigned supervisors ensured the data completeness, accuracy and consistency.

### Operational definitions

**Children**: Individuals with age less than 15 years old.

**Virological failure**: Viral load above 1000 copies/mL based on two consecutive viral load measurements in 3 months, with adherence support following the first viral load test (2).

**Immunological failure**: Fall of CD4 count to baseline (or below) or 50% fall from on treatment peak value or Persistent CD4 level below100 cells/mm3 (2).

**Clinical failure**: New or recurrent WHO stage 3 and 4 condition

**Treatment failure**: - Patients have clinical, immunological or virological failures.

**Poor Adherence:** is defined in terms of total missed appointments described by greater than three days per month (2).

**Censored**: Patients who completed their follow up, transferred out, or lost without developing virological failure.

**Time to detection of treatment failure**: The time between ART initiation and detection of treatment failure of first-line ART.

### Data processing and analysis

The collected data were entered into Epi Info version 7 and analyzed using SPSS version 21. Data cleaning was performed to check for frequencies, missed or error values, and identified errors were corrected after revision of the original completed questionnaire. Descriptive statistics was used to describe socio-demographic, clinical, laboratory and treatment-related characteristics of patients. The Kaplan–Meier method was used to estimate the probability of treatment failure at different time points. Incidence of treatment failure was calculated per person months of observation. To identify the predictors of the first line treatment failure the Cox proportional hazards model was applied. First, a bivariable analysis was conducted and variables with p-value less than 0.25 were entered into a multivariable analysis to control the effects of confounders. In the multivariable analysis, variables with p-value less than 0.05 were declared as statistically significant with 95% confidence interval.

### Ethical considerations

Ethical clearance was obtained from the Institution Review Board (IRB) of Addis Ababa Public Health Research and Emergency Management Directorate. Then, official letter was submitted to the selected health facilities and permission was obtained to access the ART client’s database and charts. To maintain confidentiality patient name and unique ART number were not included in the data extraction checklist. Furthermore, confidentiality of data was kept at all levels of the study and not used for any other purposes than the stated study objectives.

## Results

### Socio-demographic and clinical characteristics

A total of 250 medical charts were included in this study with a 100% response rate. Slightly more than half (55.2%) were female while the remaining 44.8% were male. This showed that a good proportion of male and female sample was taken for the study. The mean age at start of ART was 10.5 years (SD ± 3.36) with minimum age of 1 and maximum age of 14 years. The majority of participants (58.8%) fell in the age group 10-14 years. The majority of the participants (80%) lived with at least one of their biological parents. About three quarters (76.4%) of the care givers had a positive HIV serologic status. The mean follow-up period after ART initiation was 49.2 months ranging from 6 to 95 months (**Table 1**).

**Table 1.**
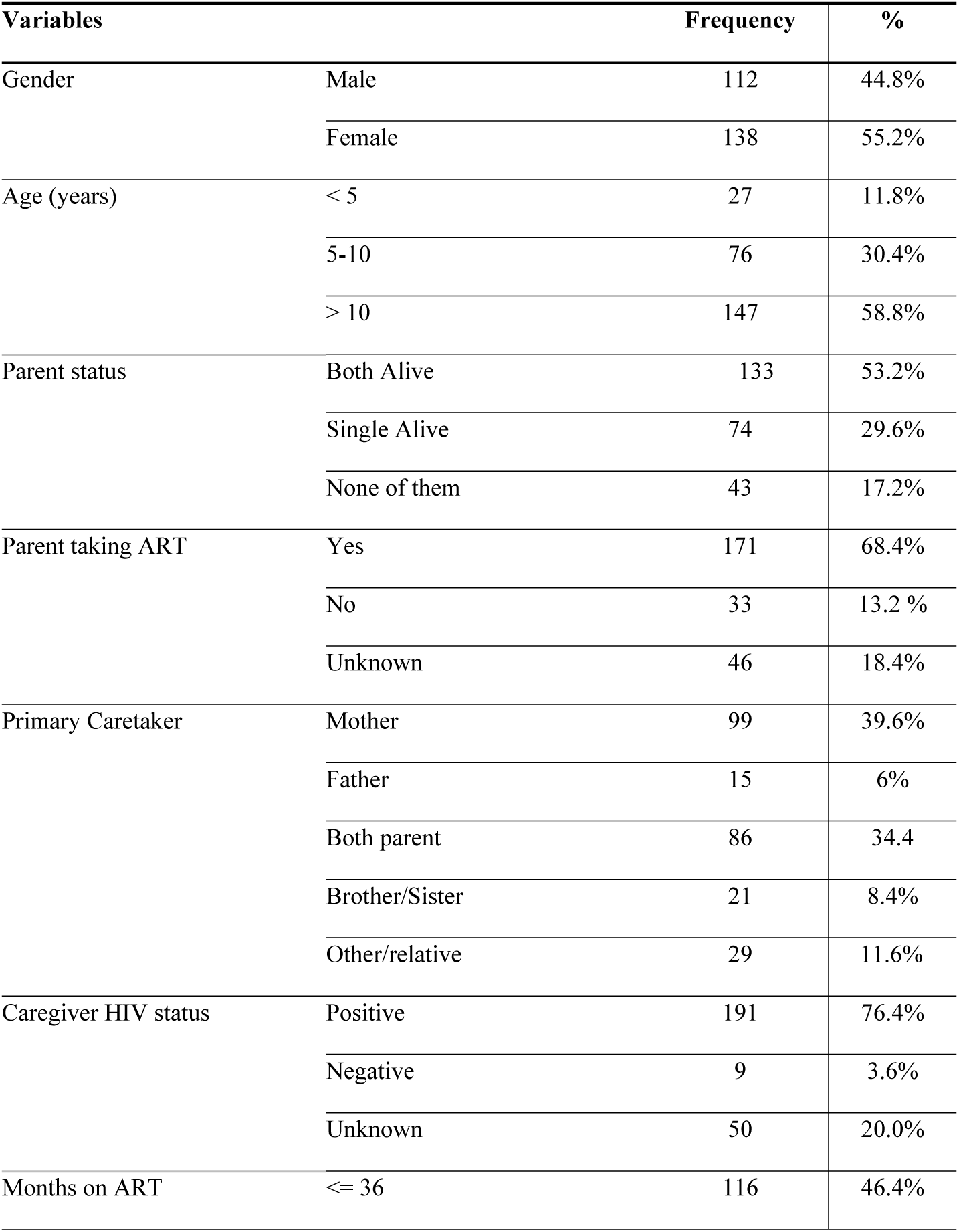
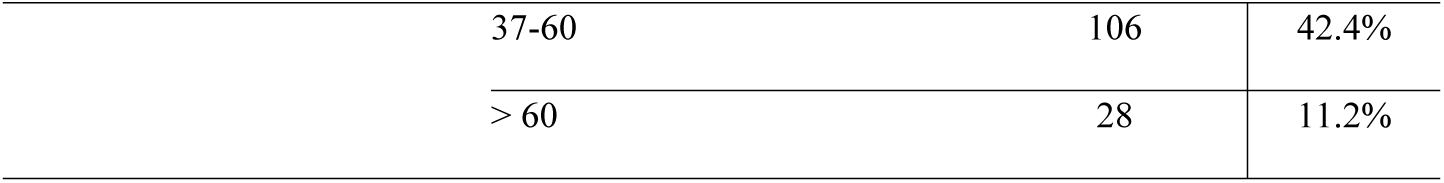
Socio-demographic characteristics children on ART at Kolfe Keranyo sub-city, Addis Ababa, Ethiopia from 2013 to 2020 (n = 250)

| **Variables** |  | **Frequency** | **%** |
| --- | --- | --- | --- |
| Gender | Male | 112 | 44.8% |
|  | Female | 138 | 55.2% |
| Age (years) | < 5 | 27 | 11.8% |
|  | 5-10 | 76 | 30.4% |
|  | > 10 | 147 | 58.8% |
| Parent status | Both Alive | 133 | 53.2% |
|  | Single Alive | 74 | 29.6% |
|  | None of them | 43 | 17.2% |
| Parent taking ART | Yes | 171 | 68.4% |
|  | No | 33 | 13.2 % |
|  | Unknown | 46 | 18.4% |
| Primary Caretaker | Mother | 99 | 39.6% |
|  | Father | 15 | 6% |
|  | Both parent | 86 | 34.4 |
|  | Brother/Sister | 21 | 8.4% |
|  | Other/relative | 29 | 11.6% |
| Caregiver HIV status | Positive | 191 | 76.4% |
|  | Negative | 9 | 3.6% |
|  | Unknown | 50 | 20.0% |
| Months on ART | <= 36 | 116 | 46.4% |

|  |  |  |
| --- | --- | --- |
| 37-60 | 106 | 42.4% |
| > 60 | 28 | 11.2% |

### Baseline clinical and antiretroviral treatment characteristics

One hundred nineteen (47.6%) of study participants were found on AZT-3TC-NVP ART regimen during ART initiation. The median baseline CD4 count was 461 cells/mm3 (IQR: 304-607). More than half (63.6%) of the study participants had experience of regimen change during their ART follow-up time when the program shifted from D4T to AZT or TDF and NNRTI to DTG or LPV/r. During their follow–up time, the majority of the study participants (88.8%) did not take PMTCT prophylaxis, even if born from HIV infected mothers. Among the children on ART, half (50.4%) took Isoniazid prophylaxis on their follow-up time (**Table 2**).

**Table 2.** Baseline clinical and antiretroviral treatment characteristics of children on first line ART at Kolfe Keranyo sub-city, Addis Ababa, Ethiopia from 2013 to 2020 (n = 250)

| Variables |  | Frequency | % |
| --- | --- | --- | --- |
| WHO stage | Stage I | 141 | 56.4% |
|  | Stage II | 65 | 26% |
|  | Stage III | 36 | 14.4% |
|  | Stage IV | 8 | 3.2% |
| Regimen at initiation | D4T/3TC/NVP | 14 | 5.6% |
|  | D4T/3TC/EFV | 2 | 0.8% |
|  | AZT/3TC/NVP | 119 | 47.6% |
|  | AZT/3TC/EFV | 52 | 20.8% |
|  | TDF/3TC/DTG | 14 | 5.6% |
|  | ABC/3TC/LPV/r | 14 | 5.6% |
|  | ABC/3TC/DTG | 7 | 2.8% |
|  | AZT/3TC/LPV/r | 6 | 2.4% |
|  | TDF/3TC/EFV | 20 | 8% |
| Infant Prophylaxis | Yes | 28 | 11.2% |
|  | No | 222 | 88.8% |
| Disclosure of HIV status | Disclosed | 116 | 46.4% |
|  | Not Disclosed | 106 | 42.4% |
|  | Unknown | 28 | 11.2% |
| Status last visit | Alive | 220 | 88% |
|  | Transfer Out | 15 | 6 % |
|  | Dead | 2 | 0.8% |
|  | Drop | 8 | 3.2% |
|  | Lost | 5 | 2% |
| Isoniazid Prophylaxis | Yes | 126 | 50.4% |
|  | No | 124 | 49.6% |
| Regimen substitutions | Yes | 159 | 63.6% |
|  | No | 91 | 36.4 % |

### Incidence of Treatment failure

Among the 250 study subjects, 82.8% were right censored (free of treatment failure). It was found that a total of 43 subjects (17.2%) have treatment failure. Of the failures, 62.8% were virological, 18.6% were immunological, and 2.3% were clinical failures. A mixed virological and immunological failure accounted for 16.3%. The overall incidence rate was 3.45 (95% CI: 2.57-4.67) per 1000 person-years of observations. The incidence of virological failure was 2.74 (95% CI: 1.95-3.83) per 1000 person-months, immunological failure was 1.13 (95% CI: 0.67-1.91) per 1000 person-months and clinical failure was 0.08 (95% CI: 0.01-21.5) per 1000 person-months (**Fig. 2)**.

**Fig. 2:**
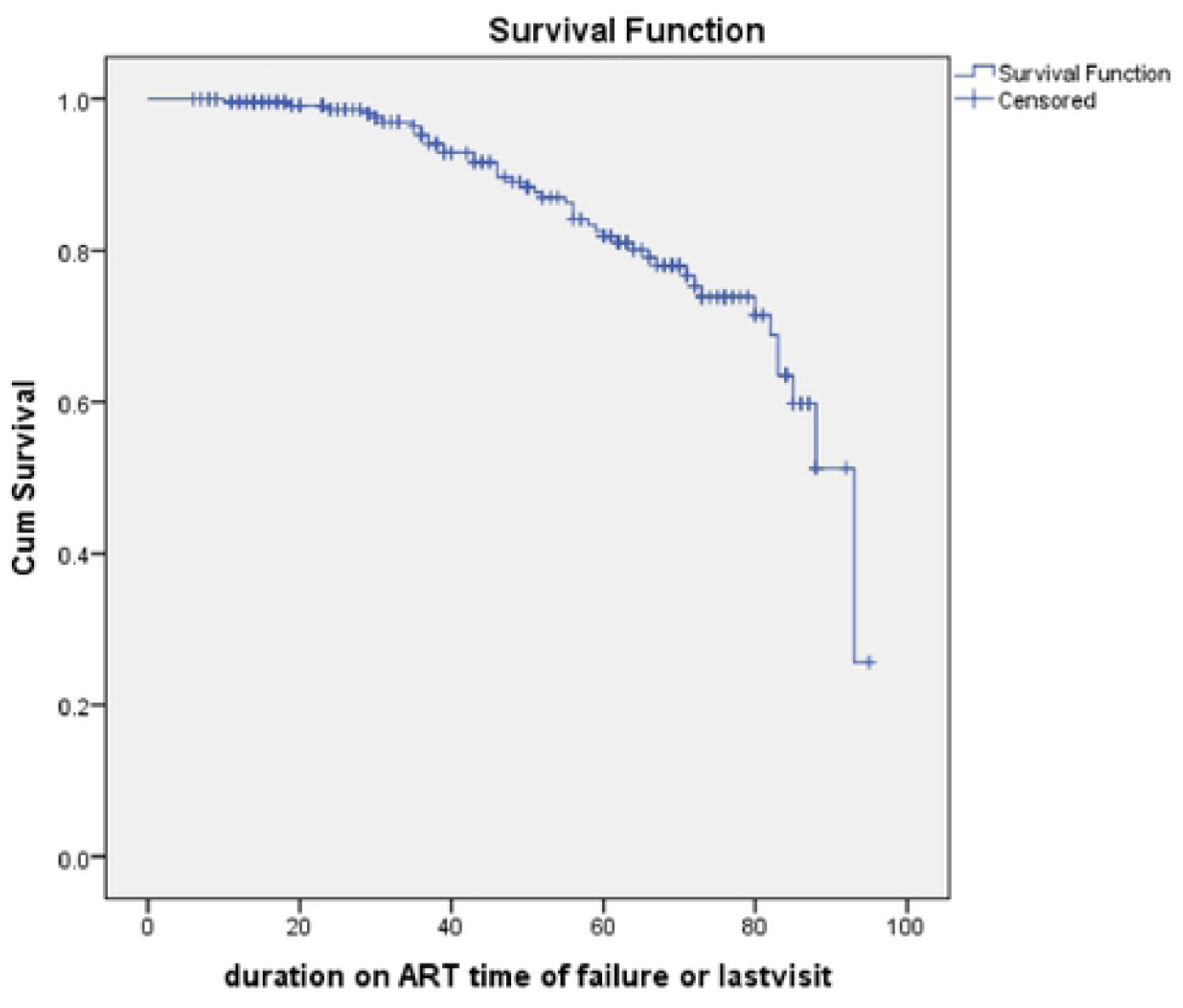
The Kaplan Meier Survival function of children on first line ART at Kolfe Keranyo sub-city, Addis Abada, Ethiopia from 2013 to 2020 (n = 250)

### Predictors of treatment failure

The bivariable Cox proportional hazard analysis showed that baseline WHO stage III and IV, regimen substitution, parent’s HIV status, infant prophylaxis at birth, initial drug regimen, parent taking ART, and episodes of poor adherence were the significant predictors of treatment failure.

In the multivariable Cox regression analysis, infant prophylaxis, drugs substitutions, initial drug regimen, and poor adherence were the predictors of treatment failure. Accordingly, children who took infant prophylaxis at birth had about 3.6 times (AHR: 3.59, 95% CI: 1.65-7,82) higher risk towards treatment failure when compared to those who didn’t took infant prophylaxis. Children with drug substitutions were by 82% less likely to suffer from treatment failure when compared to those without drug substitutions (AHR: 0.18, 95% CI: 0.09-0.37). Children who were on AZT/3TC/NVP based initial treatment regimen had about 2.3 times (AHR: 2.27, 95% CI: 1.14-4.25) higher risk towards treatment failure when compared to those who were on other initial regimens. Moreover, children with more than 3 episodes of poor ART adherence had about 2.3 times (AHR: 2.27, 95% CI: 1.17-4.38) higher risk towards treatment failure when compare to those with less than 3 episodes of poor adherence (**Table 3**).

**Table 3.**
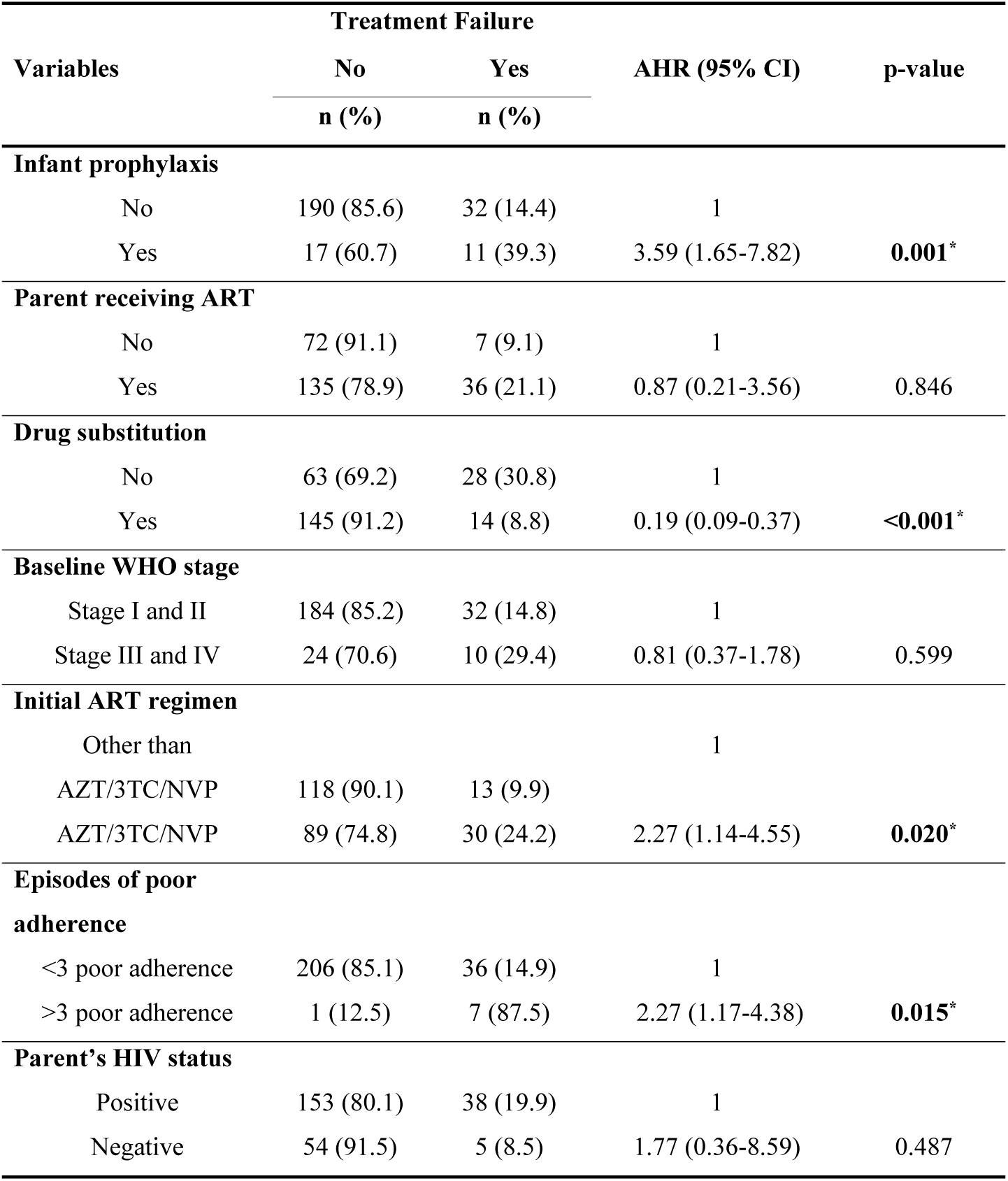
Multivariable Cox regression analysis showing predictors of ART treatment failure at Kolfe Keranyo sub-city, Addis Ababa, Ethiopia from 2013 to 2020 (n = 250)

## Discussion

Despite the availability of ART drugs, antiretroviral treatment failure has been a concerning issue globally especially in developing countries like Ethiopia (10,16). This study was conducted to assess the incidence and predictors of treatment failure among children on first line antiretroviral therapy in Kolfe Keranyo sub-city Addis Ababa, Ethiopia. In this study a 17.2% treatment failure with 3.45 (95% CI: 2.57-4.67) per 1000 person-month incidence rate were observed. Moreover, infant prophylaxis, drugs substitutions, initial drug regimen, and poor adherence were the predictors of treatment failure.

The overall treatment failure in this study was 17.2%; virological, immunological and clinical failure accounted for 79%, 18.6% and 2.3% respectively. The current finding is comparable with the finding reported at Fiche and Kuyu hospital, 18.9% (17), Gondar hospital, 18.2% (18), and a study in Ethiopia, 17.3% (15). On the other hand, treatment failure was found to be lower as compared to the findings reported in black lion hospital, 22.6% (19), Kenya, 37% (20), Uganda, 34% (9), and Zimbabwe, 30.6% (21). However, it is higher than that of the study conducted in Amhara regional state, 7.7% (8), Amhara region referral hospitals, 12.1% (1), Felegehiwot referral hospital, 10.7% (22), India, 5% (23), and Addis Ababa, 14.1% (11).

In this study, virological failure shared the highest number of failure followed by immunologic failure and only small proportion of clinical failure which is similar to the findings in Amhara region referral hospital (1), Uganda (9), and Kenya (20). The reasonfor the highest proportion of virological failure might be due to that virologic failure early detects treatment failure than immunological and clinical failure.

In this study PMTCT prophylaxis at birth was found as a predictor of treatment failure. Comparable findings was reported in studies conducted in Uganda (9) and Gondar (18). NVP is the most commonly used drugs during PMTCT and it is also used as a backbone for NNRTI regimen of HAART, which might result in a higher risk of treatment failure.

This study showed that drug substitution was about 18% protective for treatment failure. This finding contradicts with the study results in Addis Ababa (11), Gondar (18) and Felegehiwot hospital (22). This might be due to the previous use of highly mutated drugs as a substitute drug. Currently, most of substitutions are provided by newly drug available drugs (protease inhibitors and integrase inhibitors) for optimized pediatric regimen.

AZT/3TC/NVP based initial regimen was also one of the predictors of the treatment failure in this study. This is similar to the findings reported in studies conducted in Amhara region referral hospital (1) and Addis Ababa (11). Whereas, studies conducted in Uganda (9) and Zimbabwe (21) reported NVP based regimen as a predictor of treatment failure. The fact that AZT/3TC/NVP regimen is not being used currently could explain the observed variations.

Poor ART adherence among children on first-line ART was another predictor of treatment failure in this study. This finding is supported by various previous studies conducted in Ethiopia (15), Uganda (9), Gondar (18), Felegehiwot hospital (22), Amhara region referral hospital (8), and Fiche and Kuyu hospital in Oromia region(17). This could be explained by poor adherence highly contributed to mutations and resistance of antiretroviral drugs which might result in treatment failure.

Contrary to the findings in several other studies (11,17,22,24), baseline CD4 count was not a predictor of treatment failure in this study. Whereas, consistent findings were reported in studies conducted in Amhara region different hospitals (1,8). This variation could be explained by recently, ART is being provided for every HIV positive individual and baseline CD4 is not being done routinely for each and every patient. In fact, patients with a very low CD4 count are more likely to have been exposed to different OIs (opportunistic infections).

Studies conducted in Amhara region referral hospitals (1), Black lion hospital (19), and Fiche and Kuyu (17) reported that WHO clinical stages and OIs as significant predictors of treatment failure. Whereas, this study reported a contradictory finding, which is consistent with the findings reported in Uganda (9). This variation might be due to the test and treat strategy differences, there is a high public health advocacy approach to ART nowadays. Besides, the use of small sample size could also be another reason.

Moreover, HIV status disclosure and the status of primary caretaker were not predictors of treatment failure in this study. These findings were in contrast with the findings reported in Amhara region (8) and Black lion hospital (19). These variations might be due to difference in sample size. Most of the children’s caretakers were their own parents, this might also be another reason for the observed variations.

Although this is retrospective cohort study, there were few incomplete documentations that can be considered as a limitation. The temporal relationship between CD4 count determinations and concurrent illnesses was also difficult to determine due to the retrospective nature of the study.

## Conclusions

The proportion of treatment failure among children on first-line ART in Kolfe Keranyo sub-city in Addis Ababa was found to be high according to UNAIDs plan of 3^rd^ 90. Poor ART adherence, Infant prophylaxis for PMTCT, drug substitution, initial regimen, and poor adherence were found to be the predictors of first-line ART treatment failure. Hence, stakeholders working on HIV need to focus on these factors in order to minimize the rate of treatment failure. Early detection of treatment failure is important for optimal management of HIV infected children receiving ART.

Parents or caregivers should strictly support children on antiretroviral treatment adherence. As most of the children are minors and did not know their HIV status, parents or caregivers should be responsible in following children on medication adherence. Similarly, health care providers should also strictly support children on medication adherence. Effective trainings to health professionals based on revised guidelines is also essential. Furthermore, future studies should examine other underlying relevant factors including viral load as a measurement for treatment failure.

## Data Availability

All relevant data are within the manuscript and its Supporting Information files.

## Acknowledgements

We would also like to express our gratitude to Addis Ababa City Administration Health Bureau and Kolfe Keranyo sub-city for allowing us to access the required patient data. We also would like to thank the data collectors and supervisors who contributed significantly for the success of this research work.

## Authors’ Contributions

**Conceptualization:** Meseret Misasew.

**Data Curation:** Meseret Misasew.

**Formal Analysis:** Meseret Misasew, Daniel Angassa.

**Funding Acquisition:** Meseret Misasew.

**Investigation:** Meseret Misasew, Daniel Angassa.

**Methodology:** Meseret Misasew, Daniel Angassa.

**Project Administration:** Meseret Misasew.

**Resources:** Meseret Misasew.

**Software:** Meseret Misasew.

**Supervision:** Meseret Misasew.

**Validation:** Meseret Misasew, Daniel Angassa.

**Visualization:** Meseret Misasew.

**Writing – Original Draft Preparation:** Meseret Misasew, Daniel Angassa.

**Writing – Review & Editing:** Meseret Misasew, Daniel Angassa.

## Abbreviations

AIDS: Acquired Immune Deficiency Syndrome
ART: Antiretroviral Treatment
AZT: Zidovudine
DTG: Dolutegravir
HAART: Highly Active Antiretroviral Therapy
HIV: Human Immunodeficiency Virus
IRB: Institution Review Board
LPV/r: Lopinavir / Ritonavir
NNRTI: Non-Nucleotide Reverse Transcriptase Inhibitor
NRTI: Nucleotide Reverses Transcriptase Inhibitor
NVP: Nevirapine
OI: Opportunistic Infection
PI: Protease Inhibiter
PMTCT: Prevention of Mother to Child Transmission
SPSS: Statistical Package for Social Sciences
TB: Tuberculosis
VL: Viral Load
WHO: World Health Organization
3TC: Lamivudine

## Availability of data and materials

The datasets used and/or analyzed during the current study are available from the corresponding author on reasonable request.

